# Incidence and risk factors of omicron variant SARS-CoV-2 breakthrough infection among vaccinated and boosted individuals

**DOI:** 10.1101/2024.04.03.24305293

**Authors:** Fabiola M. Moreno-Echevarria, Mathew T. Caputo, Daniel M. Camp, Susheel Reddy, Chad J. Achenbach

**Author notes:** Corresponding author: Chad J. Achenbach, M.D., M.P.H., Havey Institute for Global Health, Feinberg School of Medicine, Northwestern University, 710 N Lake Shore Drive, Chicago, IL 60611. These authors contributed equally to this work (Fabiola M. Moreno-Echevarria, Mathew T. Caputo, Chad J. Achenbach). These authors also contributed equally to this work (Daniel M. Camp, Susheel Reddy).

## Abstract

**Background:** SARS-CoV-2 vaccines have been shown to be safe and effective against infection and severe COVID-19 disease worldwide. Certain co-morbid conditions cause immune dysfunction and may reduce immune response to vaccination. In contrast, those with co-morbidities may practice infection prevention strategies. Thus, the real-world clinical impact of co-morbidities on SARS-CoV-2 infection in the recent post-vaccination period is not well established. We performed this study to understand the epidemiology of Omicron breakthrough infection and evaluate associations with number of comorbidities in a vaccinated and boosted population.

**Methods and Findings:** We performed a retrospective clinical cohort study utilizing the Northwestern Medicine Enterprise Data Warehouse. Our study population was identified as fully vaccinated adults with at least one booster. The primary risk factor of interest was the number of co-morbidities. Our primary outcome was incidence and time to first positive SARS-CoV-2 molecular test in the Omicron predominant era. We performed multivariable analyses stratified by calendar time using Cox modeling to determine hazard of SARS-CoV-2. In total, 133,191 patients were analyzed. Having 3+ comorbidities was associated with increased hazard for breakthrough (HR=1.2 CI 1.2-1.6). During the second half of the study, having 2 comorbidities (HR= 1.1 95% CI 1.02-1.2) and having 3+ comorbidities (HR 1.7, 95% CI 1.5-1.9) were associated with increased hazard for Omicron breakthrough. Older age was associated with decreased hazard in the first 6 months of follow-up. Interaction terms for calendar time indicated significant changes in hazard for many factors between the first and second halves of the follow-up period.

**Conclusions:** Omicron breakthrough is common with significantly higher risk for our most vulnerable patients with multiple co-morbidities. Age related behavioral factors play an important role in breakthrough infection with the highest incidence among young adults. Our findings reflect real-world differences in immunity and exposure risk behaviors for populations vulnerable to COVID-19.

## Introduction

Vaccines against SARS-COV-2, have been developed and shown in numerous studies to be safe and highly effective at reducing SARS-CoV-2 infection and COVID-19 disease [1–3]. However, clinical trials and population-based observational studies excluded or did not compare certain groups at highest risk for severe outcomes of COVID-19. The impact of age and burden of immune disorders or certain chronic conditions on vaccine effectiveness (including boosting) in terms of acquisition of infection, has not been adequately studied in our current era dominated by omicron SARS-CoV-2 subvariants.

Although there have been impressive advances in our understanding of protective immunity against COVID-19 after vaccination and natural infection, risk of infection is not completely understood for our most vulnerable patients including individuals who are immune compromised (age, HIV, malignancies, solid organ transplant, stem cell transplant) or have chronic illnesses (diabetes, obesity, chronic liver disease, and chronic kidney disease). SARS-CoV-2 vaccines stimulate both B and T cell responses to virus spike protein to elicit an effective immune response [4]. Those with dysfunctional immunity have been observed to have lower responses to vaccination with antibody titers as indicators of immunogenicity [5]. Questions remain about how lower immunogenicity translates into diminished clinical effectiveness of COVID-19 vaccines in real world populations with different co-morbidities. Individuals who have chronic disease, advanced age, and/or immunodeficiencies may be at higher risk for breakthrough infection due to poor vaccine response (defined as COVID-19 infection after completion of all require doses with a typical 2-week lag period) [6]; however, they may also practice better infection prevention such as mask wearing, avoiding travel or large gatherings, and social distancing [7].

The incidence of SARS-CoV-2 breakthrough infection is an increasingly important issue worldwide, with vulnerable populations at high risk of infection at a time when vaccine-induced immunity may not be fully optimized. To gain insight into this issue and to inform public health decision making, our study aimed and was able to determine the incidence and risk factors associated with SARS-CoV-2 breakthrough infection in the Omicron-variant era among vaccinated and boosted individuals.

## Methods

### Study Design

We performed a clinical cohort study of breakthrough infection in the first year of SARS-CoV-2 Omicron-variant era (January 1, 2022, until December 31, 2022) among fully vaccinated and boosted adults (18 years and older) as per CDC/FDA vaccine guidelines for COVID-19 [8], All demographic, lab, vaccine, and comorbidity data were collected from the Northwestern Medicine (NM) Enterprise Data Warehouse (EDW)[9]. Each participant had a unique study identifier and protected health information was stored separately with access limited to the principal investigator.

### Population & Definitions

We included adults that were boosted prior to the Omicron era, which we defined as those who received at least 3 (first dose mRNA) or 2 (first dose J&J) SARS-CoV-2 vaccine doses prior to December 15, 2021. To include a representative population that was likely to have SARS-CoV-2 testing performed within the NM system, we only included patients if they had at least two medical system visits (including inpatient, outpatient, telemedicine, and lab testing) at least 180 days apart between January 1, 2020, and December 15, 2021.

We observed individuals in this cohort from January 1, 2022, to December 31, 2022, for incident breakthrough infection with SARS-CoV-2 as our primary outcome defined as the first positive SARS-CoV-2 PCR test performed at an NM facility after January 1, 2022. Patients who tested positive for SARS-CoV-2 after their most recent booster dose but prior to January 1, 2022, were excluded. Data was accessed throughout this study period. Comorbidities of interest (diabetes, obesity, solid organ transplant, stem cell transplant, HIV, hematologic malignancy, chronic liver disease, and chronic kidney disease) were identified using ICD9/ICD10 coding.

### Statistical Analysis

Descriptive statistics, including median (IQR) and counts (%), were calculated for patient characteristics and compared between those with and without breakthrough SARS-CoV-2 infection during the study period. We calculated cumulative incidence as the number of breakthrough infections divided by the total number of those at risk (no prior breakthrough infection during study and not right censored) and plotted these curves with 95% confidence intervals assuming normality over the study period. Patients were right censored in cases of death, additional SARS-CoV-2 vaccines, or loss to follow-up. Loss to follow-up was defined as 90 days without a visit at NM (outpatient visit, hospital admission, or laboratory testing). We performed Cox regression modeling to determine proportional hazards of breakthrough infection with the following covariates: age, sex, race, ethnicity, time from booster dose to study period start, and number of comorbidities. We included the overall number of comorbidities rather than the specific comorbidities themselves as this study aimed to measure associations with overall health, not individual disease epidemiology. The proportional hazards assumption was assessed graphically with cumulative incidence curves for each covariate. For covariates that violated this assumption, interaction terms for calendar time were considered. Multicollinearity was assessed with generalized variance inflation factors (GVIFs). If (GVIF^1/(2⋅DF)^)^2^ > 3, removal of those variables from the model was considered. Linearity of log hazards for continuous covariates was examined with martingale residuals from the fitted model. The *car, survival* and *tidycmprsk* packages in R 4.2.3 software were used for statistical analysis and plot production[10–13].

## Results

Clinical and demographic characteristics of the cohort included and analyzed are presented in **Table 1**. In total, there were 133,191 patients in the cohort with a median (IQR) age of 61 years (47, 72) 63% female sex, 84% white, 95 % Non-Hispanic or Latino ethnicity, and 77% with any comorbid condition.

**Table 1:**
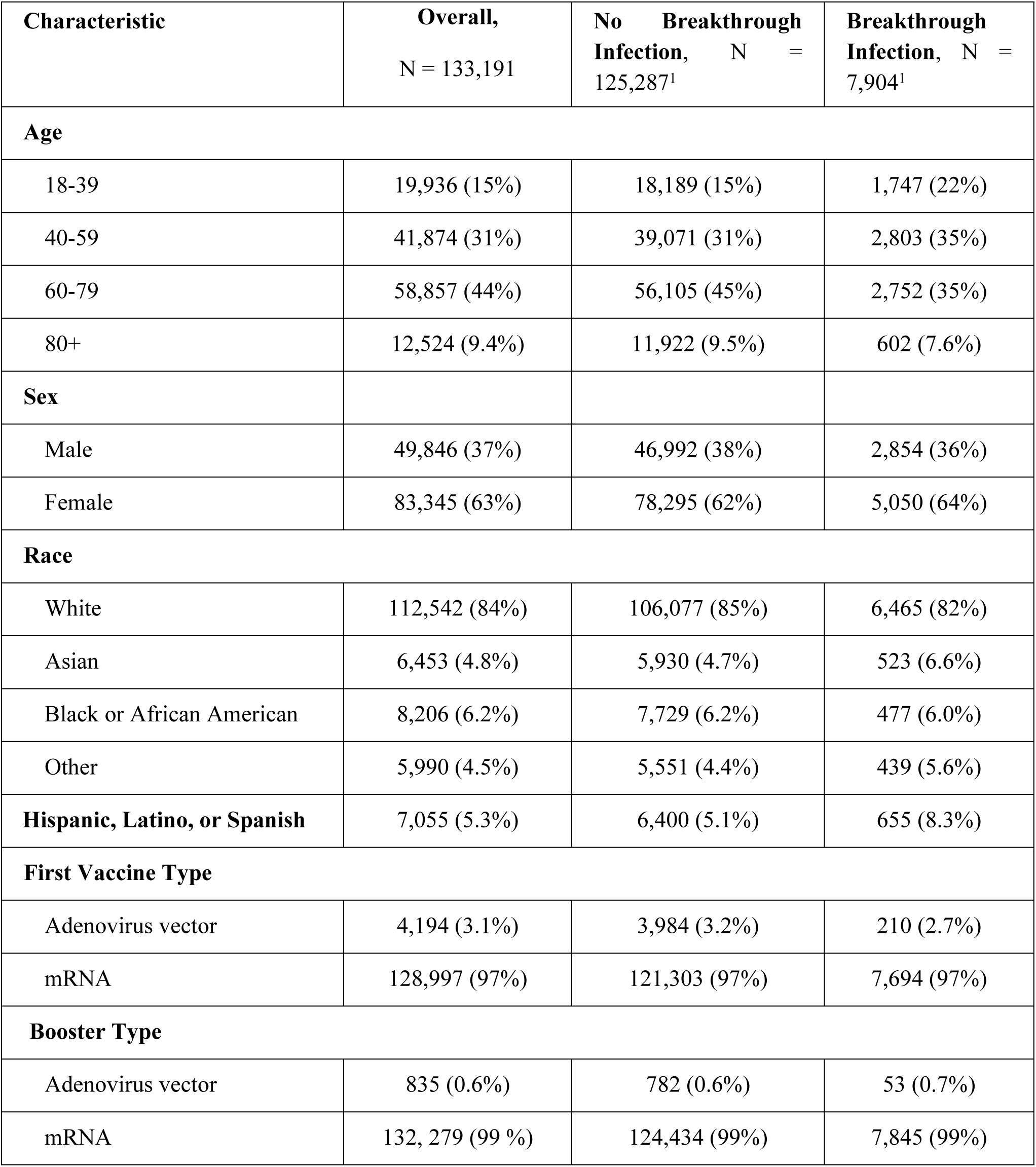
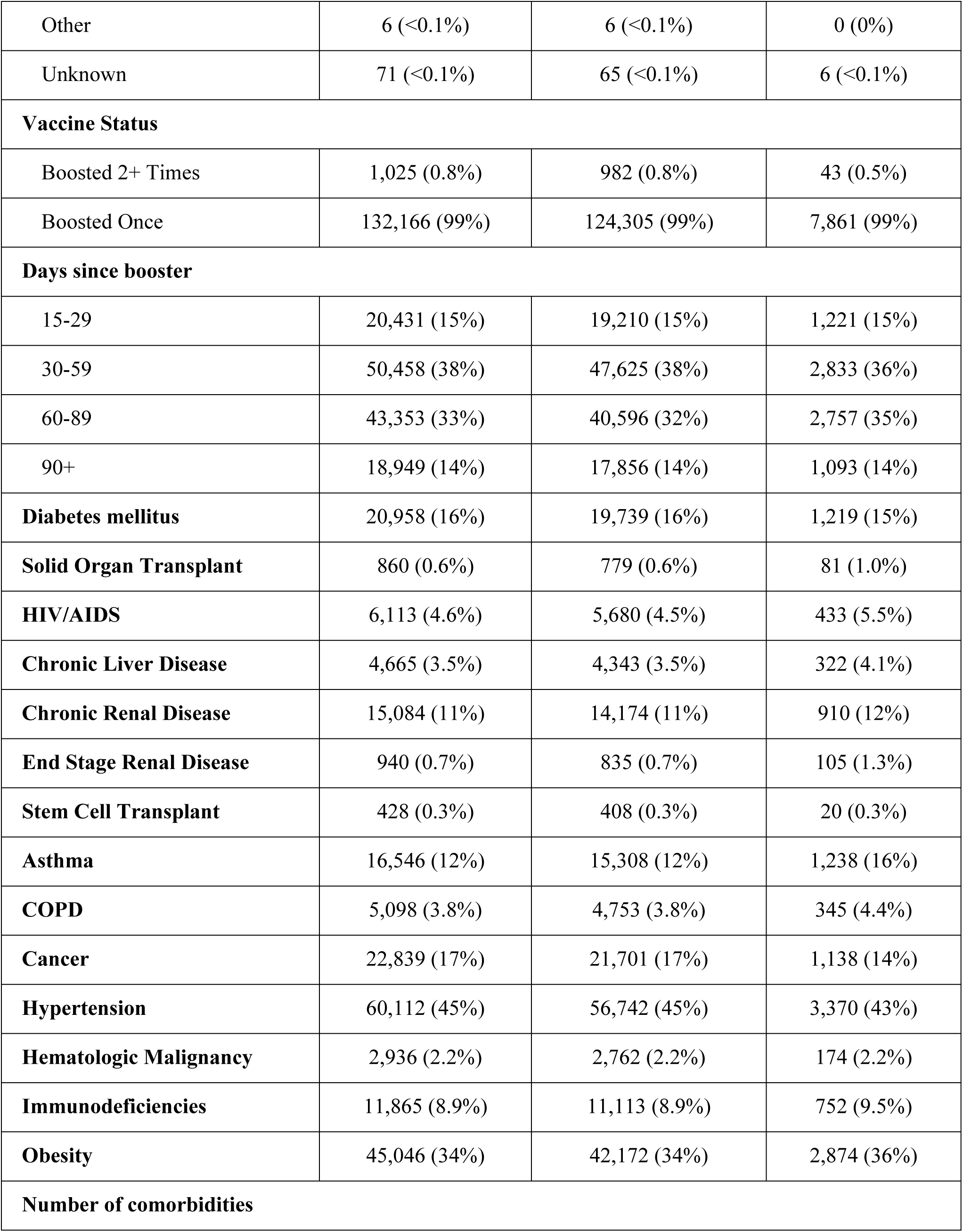
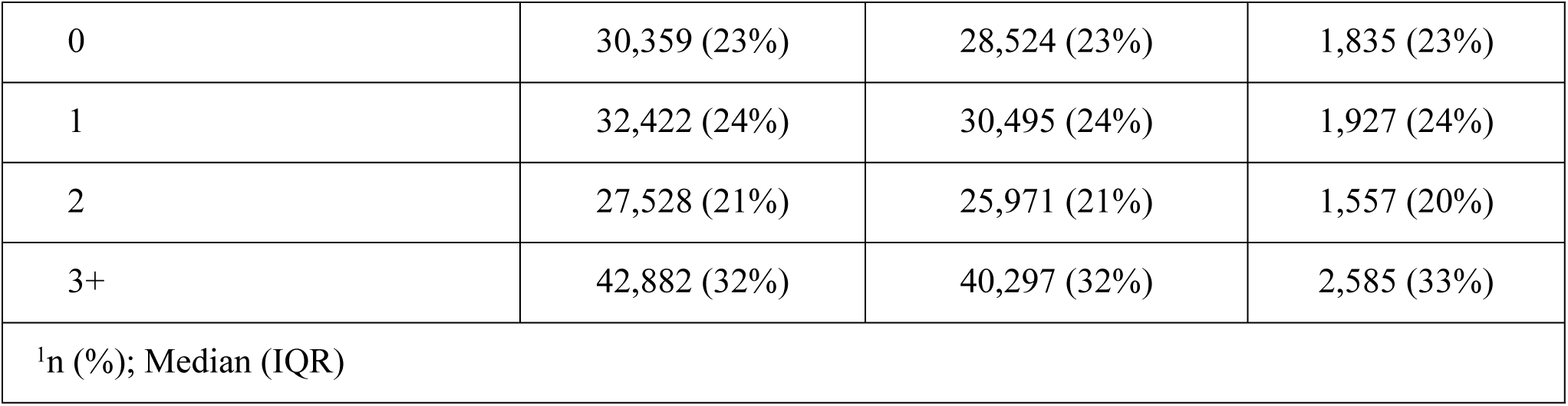
Characteristics of patients overall, with and without breakthrough SARS-CoV-2 infection.

Overall, 99% of individuals received mRNA SARS-CoV-2 booster. One booster dose vaccine was administered to 99.2% of individuals and 0.8% had received more than one booster dose.

Of the total cohort, 7,904 (5.9%) tested positive for SARS-Cov-2 infection by PCR during the study period in a median (IQR) of 135 (34, 196) days. The infections occurred in a median (IQR) of 196 (115, 260) days after a booster dose of vaccine. Approximately 31% of the total cohort was right-censored due to loss to follow-up, 21% due to additional boosters, and 0.4% due to death.

The proportional hazards assumption in Cox regression was violated for the following covariates: age, race, days since vaccination, and number of comorbidities. Graphical assessment indicated that significant changes in incidence rate trends began near the 6-month (halfway) mark of the study period. To account for this, we introduced an interaction term to calculate separate hazard ratios for periods 1 (Jan 1, 2022, to June 30, 2022) and 2 (July 1, 2022, to Dec 31, 2022) for those 4 covariates.

In Cox regression multivariable analysis **(Table 2),** increasing age was associated with reduced hazard during this first period when controlling for other covariates. The greatest reduction in hazard compared to the reference age group (18-39) was seen in the 80+ age group (HR 0.41 95% CI 0.36-0.46). Similar but lower reductions were observed in the 60-79 (HR 0.44 95% CI 0.4-0.48) and 40-59 (HR 0.71 95% CI 0.66-0.76) age groups. A significant interaction between calendar time and the hazards for age was found in each age group compared to the reference group (p < .001 for each). In the second half of the study period, only the 60-79 age group demonstrated a significantly different hazard than the reference group (HR 0.86, 95% CI 0.76-0.99). Trends in cumulative incidence stratified by number of comorbidities in are observed in **Fig. 1**.

**Fig. 1.**
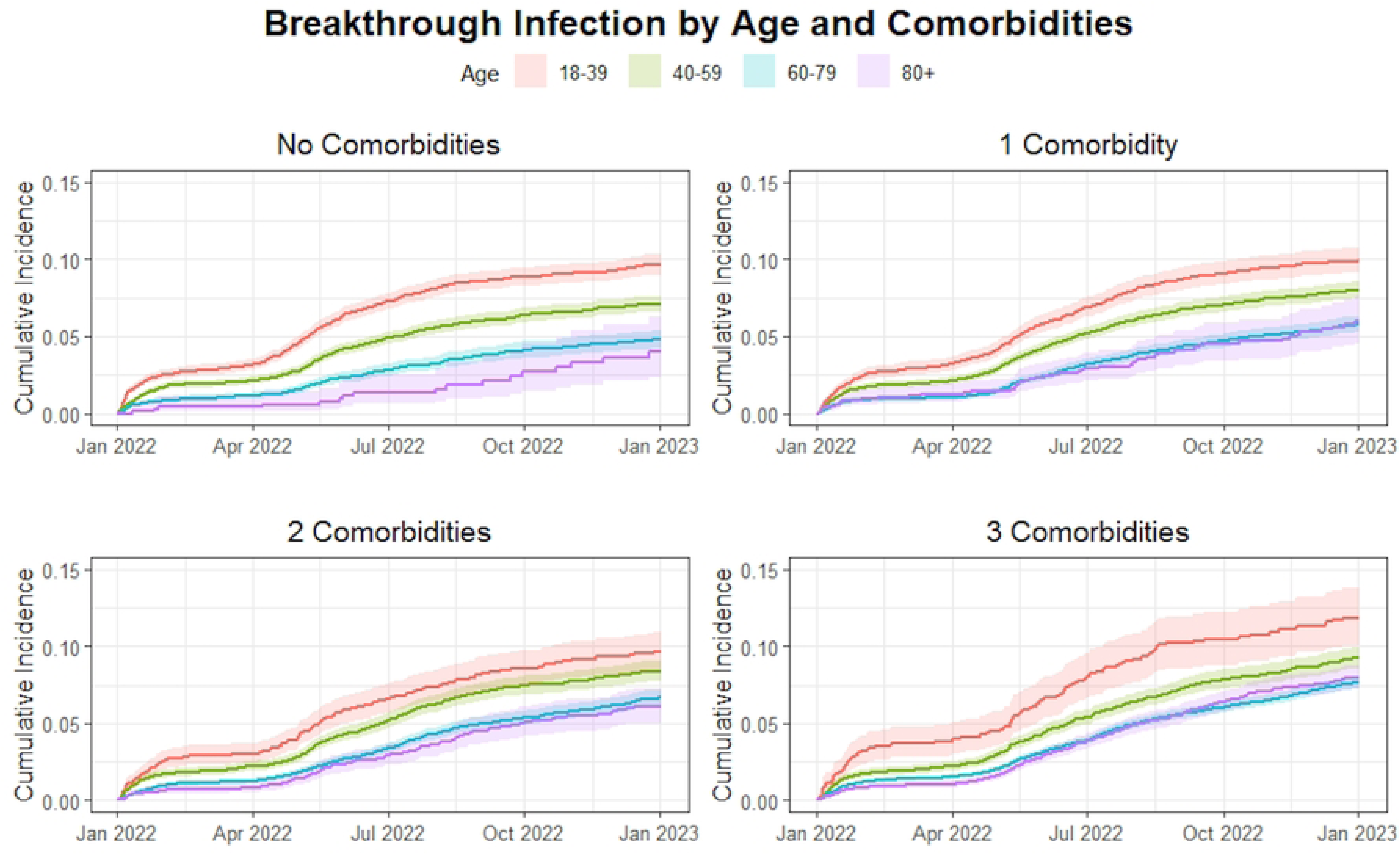
Cumulative incidence curve for COVID-19 breakthrough infection by age when stratified by number of comorbidities (per panel).

**Fig. 2.**
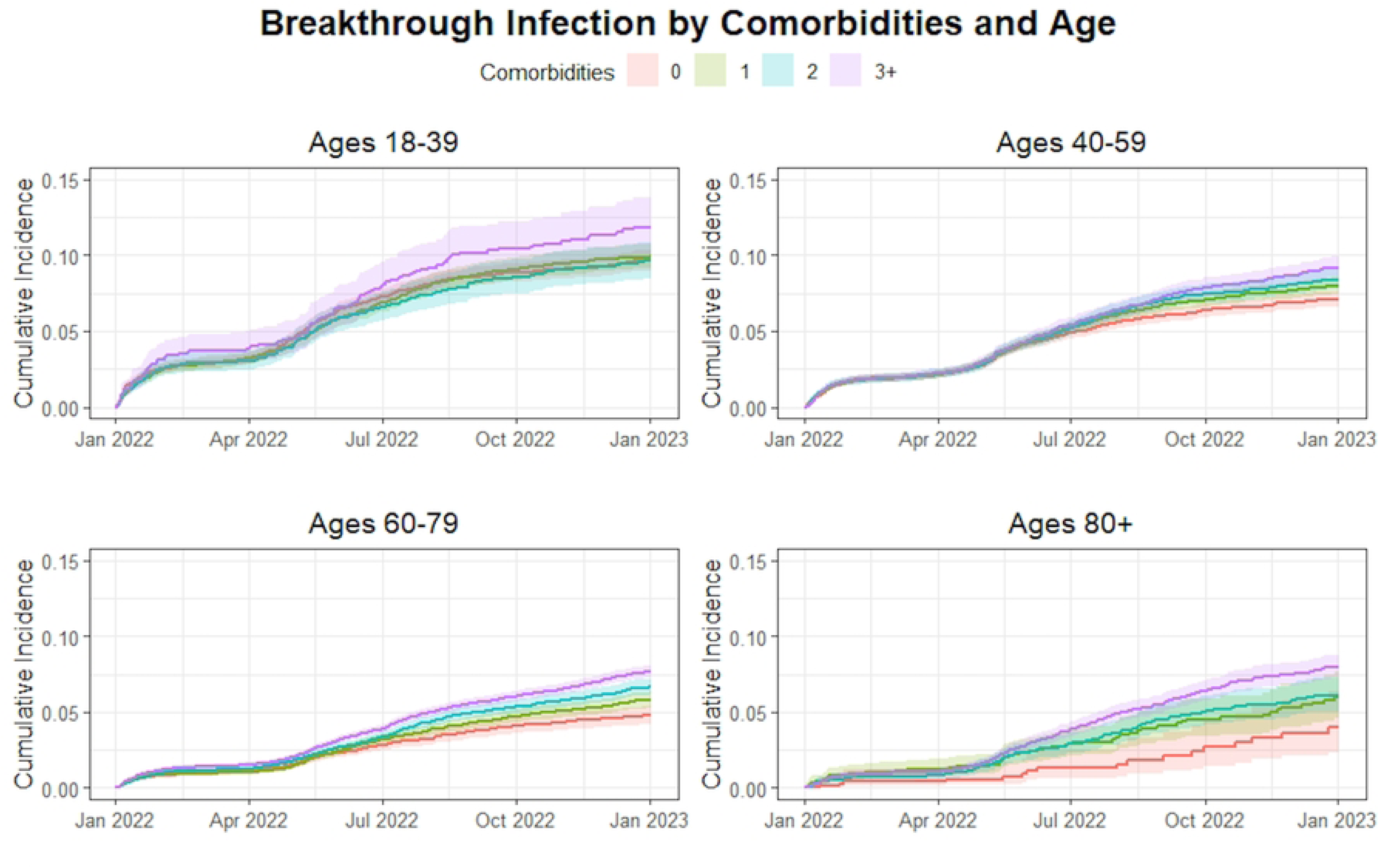
Cumulative incidence curve for COVID-19 breakthrough infection by number of comorbidities when stratified by age category (per panel).

**Table 2:** Multivariable analysis for hazard of breakthrough SARS-CoV-2 infection using Cox modeling.

| Covariate | Hazard Ratio (95% CI),<br>Period 1 | Hazard Ratio (95% CI),<br>Period 2 | Hazard Ratio (95% CI),<br>Both Periods | Covariate x<br>Period<br>Interaction<br>Term Type 3<br>p-value |
| --- | --- | --- | --- | --- |
| **Age Group** |  |  |  |  |
| 18-39 | Ref | Ref | Ref |  |
| 40-59 | 0.712 (0.664, 0.763) * | 0.921 (0.809, 1.049) | 0.751 (0.706, 0.798) * | < .001 |
| 60-79 | 0.442 (0.409, 0.477) * | 0.863 (0.756, 0.985) * | 0.529 (0.496, 0.565) * | < .001 |
| 80+ | 0.406 (0.359, 0.459) * | 0.953 (0.800, 1.136) | 0.522 (0.472, 0.576) * | < .001 |
| **Male** | Ref | Ref | Ref |  |
| **Female** |  |  | 1.010 (0.964, 1.058) |  |
| **Race** |  |  |  |  |
| White | Ref | Ref | Ref |  |
| Asian | 1.320 (1.190, 1.463) * | 1.232 (1.028, 1.476) * | 1.305 (1.193, 1.428) | .517 |
| Black or African<br>American | 0.828 (0.734, 0.935) * | 1.164 (1.003, 1.351) * | 0.945 (0.861, 1.038) | < .001 |
| Other | 0.944 (0.836, 1.068) | 0.987 (0.816, 1.193) | 0.959 (0.862, 1.067) | .691 |
| **Not Hispanic,**<br>**Latino, or**<br>**Spanish** |  |  | Ref |  |
| **Hispanic, Latino,**<br>**or Spanish** |  |  | 1.480 (1.354, 1.618) * |  |
| **Weeks from**<br>**Booster to study**<br>**start** | 1.034 (1.027, 1.040) * | 1.013 (1.002, 1.023) * | 1.028 (1.023, 1.033) * | < .001 |
| **Number of Comorbidities** |  |  |  |  |
| 0 | Ref | Ref | Ref |  |
| 1 | 1.009 (0.938, 1.087) | 1.271 (1.110, 1.455) * | 1.057 (0.991, 1.127) | .003 |
| 2 | 1.022 (0.940, 1.109) | 1.449 (1.260, 1.665) * | 1.108 (1.033, 1.189) * | < .001 |
| 3+ | 1.163 (1.075, 1.258) * | 1.726 (1.513, 1.969) * | 1.288 (1.205, 1.377) * | < .001 |
| *p < .05 |  |  |  |  |
| Period 1 = January 1, 2022, to June 30, 2022, Period 2 = July 1, 2022, to Dec 31, 2022 |  |  |  |  |

Cox regression modeling (**Table 2**) also revealed an increased hazard for those of Asian race compared to the reference group (white race) in both periods (period 1: HR 1.32 95% CI 1.19-1.46; period 2 HR 1.23 95% CI 1.03-1.48). Significant associations with hazard were seen in the Black or African American race group compared to the reference group, though the directions of these associations differed between study periods (period 1: HR 0.83 95% CI 0.73-0.94; period 2: HR 1.16 95% CI 1.03-1.48). People of Hispanic, Latino, or Spanish ethnicity also had a higher hazard for COVID-19 breakthrough infection compared to those not in that ethnic group (HR 1.48 95% CI 1.35-1.62). Days from booster to start of study showed an increased hazard of 1.03 per week (95% CI 1.03-1.04) and later 1.01 per week (95% CI 1.00-1.02). The interaction term for calendar time and this variable was significant (p < .001).

Compared to having none, having one or two comorbidities was not significantly associated with increased hazard for Omicron infection during period 1 when controlling for other variables. In contrast, having three or more comorbidities was associated with increased hazard for breakthrough (HR=1.16 CI 1.08-1.26). During the second half of the study, having one comorbidity (HR = 1.27 95% CI 1.11-1.46), having two comorbidities (HR= 1.45 95% CI 1.26-1.67) and having three or more comorbidities (HR 1.73, 95% CI 1.51-1.97) were all associated with increased hazard for Omicron breakthrough compared to those with no comorbidities. Cumulative incidence over time between co-morbidity groups and stratified by age category are presented in **Fig. 2**. Cumulative incidence over time between age categories and stratified by co-morbidities groups are presented in **Fig. 1**.

## Discussion

In this large cohort study across an urban healthcare system in Chicago, we found that omicron breakthrough infections were common with a one-year cumulative incidence of 5.1% for all vaccinated and boosted individuals. We observed a higher incidence of breakthrough infections among individuals with multiple comorbidities and those who had a longer time since vaccine booster. However, age was also an important factor with increasing age independently associated with reduced risk of Omicron breakthrough likely due to less risk taking and lower exposure among the highest age groups. We found racial and Ethnic differences in infection incidence as demonstrated by increased risk of Omicron breakthrough for individuals identifying as Asian race or Hispanic/Latino ethnicity, again likely due to differences in exposure from social behaviors rather than inherent immunogenetic differences. Perhaps our most intriguing finding was that across all age groups, those with three or more co-morbidities had the highest incidence of SARS-CoV-2 infection, and this factor did not appear to alter individual behavior to lower exposure as much as age. The immune effects of co-morbidity might have overwhelmed any decrease in social behavior risk related exposure. Thus, acquisition of Omicron SARS-CoV-2 was complex with behavioral and individual risk or immunologic factors driving SARS-CoV-2 infection despite vaccination.

Our findings are consistent with previous studies that have reported a significant burden of breakthrough infections despite vaccination efforts. For example, a study by Stouten et al. [14] found a similar cumulative incidence of 11.2% among vaccinated individuals in a different urban setting. Similarly, Sun et al.[15] reported an incidence of 7.1% in those without immune dysfunction and slightly higher rates for those with specific diseases affecting the immune system. Our cumulative incidence is lower than the ones reported in these studies. This likely relates to several factors including study population, methodologic differences, and SAR-CoV-2 variant circulating during the study period. The predominant circulating variant of concern in prior studies was the Delta variant, which has been the most studied in terms of breakthrough and consistently demonstrated high incidence rates [16]. Additionally, studies conducted in rural populations have demonstrated lower incidence of breakthrough [17, 18]. One study in a small cohort in New York City reported an incidence <1% [19], but this discrepancy is likely due to study time period and significantly smaller sample size compared to our study and others with higher incidence. As expected, and generally observed, evidence suggests that breakthrough infections are common across many different populations and healthcare settings and may vary with overtime with shifts in vaccine coverage, population immunity, and circulating SARS-CoV-2 variant strain.

Our study revealed an intriguing finding in that having three or more comorbid diseases heightened the risk of breakthrough infections across all age groups. Having more comorbidities increases risk of infection greater than one would expect of the individual comorbidities. Surprisingly, this heightened risk did not seem to prompt significant alterations in individual behavior. It is plausible that the impact of these comorbidities on immunity overshadowed any potential behavioral changes. Alternatively, it could be that comorbidities are not as influential in driving behavioral and lifestyle changes as age. However, the latter would contrast with prior studies suggesting that perceived vulnerability and severity, along with self-efficacy and intention, are drivers of COVID-19 protective behaviors[20–23]. Our findings align with those of Smits et al. [24], who examined four specific comorbidities and observed that patients with two or more of these conditions also faced a greater risk than would be expected based on the individual effects of each comorbidity. This contrasts with the findings of Walmsley et al. [25], who did not identify significant variations in the rate of breakthrough infections among individuals with underlying comorbidities. Theses investigators acknowledged that this discrepancy may be due to the low number of participants with comorbid diseases in their study, which could have hindered the identification of a clear association.

Immune compromising co-morbidities have been shown to increase risk of SARS-CoV-2 infection [26];[27–29], however, we did not find any significant increased risk among those in our immunocompromised groups of stem cell or solid organ transplant recipients as has been previously reported[28]. Whether due to the impact on the immune system or lack of an effect on behaviors, our findings reveal that independent of age or type of comorbidity, an increasing amount of comorbidity burden increased the risk for Omicron SARS-CoV-2 infection after vaccination within a population most vulnerable to worse outcomes and severe COVID-19 disease. Thus, public health messing should continue to emphasize the importance of infection prevention measures within this key group.

We noted that older individuals became at higher risk for infection at later time points, suggesting reduction in vaccine response or increase in risk taking behaviors over time. This finding is consistent with a previous study that demonstrated an inverse association between age and antibody titers only three months after mRNA vaccination [30]. Similarly, a study focusing on immunocompromised individuals found that although vaccination was associated with a modest risk reduction, older individuals continued to have higher rates of breakthrough SARS-CoV-2 infections [15]. Research has shown that as the immune system ages, the number of naïve T and B cells decreases, which can lead to reduced vaccine efficiency and a predisposition to breakthrough infections. While the quality of antibodies remains unaffected, aging is associated with a decrease in the quantity of antibodies produced after vaccination, rendering individuals more susceptible to infection [31, 32]. Our results support prior evidence establishing that, independent of comorbidities or immunocompromised status, increased age is a strong risk factor for breakthrough infection likely due to declining immunity over time. Overall, this evidence suggest that the diminished quantity of antibodies produced in older individuals puts them at greater risk compared to the longer-lasting protection seen in younger people. Our findings also support recent recommendations for SARS-CoV-2 vaccine boosting approximately twice per year among individuals over 65 years of age [33].

In contrast to our findings regarding older individuals, multiple studies have identified younger age as a risk factor for COVID-19 breakthrough infection[14, 25, 34] . It’s important to note that these studies varied in design, cohort size, and follow-up duration, which may have influenced their ability to capture current evolving trends. These findings align with our observations shortly after vaccination, where younger age emerged as an important risk factor. We attribute this phenomenon to social behaviors playing a larger role than age during earlier time periods, when immunity has not yet significantly declined and vaccine response more protective. Young adults engage in more social interactions and risk-taking behaviors for respiratory virus acquisition compared to the elderly, potentially increasing their exposure to SARS-CoV-2through daily activities [35, 36]. Certain occupations, such as healthcare work, transit operation, and retail roles, have been identified as particularly high-risk for breakthrough infection despite full vaccination [37, 38]. Additionally, younger individuals may perceive themselves as less vulnerable to severe COVID-19 disease and therefore be more likely to disregard public health precautions such as social distancing and mask-wearing, or they may be less adept at recognizing the signs and symptoms of the virus, leading to further spread of the disease [39, 40]. Thus, it appears that prior to a significant decline in immunity and vaccine efficacy, behaviors among younger individuals likely drive and risk for breakthrough SARS-Co-V2 infections.

Time since last booster dose and certain racial/ethnic group also emerged as independent risk factors. A study described a similar finding in relation to the time since second vaccine dose [41]. This finding is consistent with waning vaccine immunity that has been previously described [42–46]. Our study also highlights the increased risk of Omicron SARS-CoV-2 infection among vaccinated individuals from Hispanic, Asian, and Black ethnic/racial groups. We believe it is unlikely that there is a biological basis to these associations– rather due to well-established socioeconomic factors known to play significant roles in influencing the occurrence and outcomes of SARS-CoV-2 and COVID disease [47–49]. Prior studies examining breakthrough SARS-CoV-2 infection do not comment on demographic information, likely limited geographical regions being studied. [14, 16]. However, reports in immunocompromised populations did not find any significant racial or ethnic differences [15, 50]. Disparities in healthcare access and quality may contribute to certain demographic groups facing a higher risk of SARS-CoV-2 infection[47, 51] .

Engagement in higher-risk occupations, larger household size, lower income level, distrust in healthcare, lack of health insurance, and unequal access to healthcare services are among the key contributors to these disparities [52]. Efforts to address these disparities and improve healthcare outcomes for high-risk demographic groups are needed and should include interventions aimed at expanding access to healthcare and insurance, establishing more equitable care models, and addressing the underlying social determinants of health.

Our study has several limitations that should be considered when interpreting the results. Firstly, our research was conducted using electronic medical records within a single urban healthcare system, which likely did not capture all breakthrough SARS-CoV-2 infections. Patients who sought care outside of our NM system or opted for at-home testing were not always identified in our analysis, potentially leading to an underestimation of the true cumulative incidence of breakthrough SARS-CoV-2 infections. Additionally, mild, or asymptomatic breakthrough infections may be underreported in our study, as individuals with less severe symptoms might be less likely to seek medical care or testing. Secondly, given our study population selection criteria (see above), we may have included individuals who were generally more proactive about seeking healthcare services and thus may be overrepresented. This may affect the generalizability of our findings to a broader less engaged population. Additionally, the findings of this study may not be generalizable to other populations or healthcare settings. Factors such as population demographics, vaccination rates, and healthcare infrastructure can vary widely between different regions and may impact the incidence and risk factors for breakthrough SARS-CoV-2 infections. Lastly, our study was conducted during a specific timeframe and primarily focused on the Omicron variant. The future incidence and risk factors for breakthrough SARS-Co-2 infections will likely vary with evolving population immunity (natural and/or updated vaccines) and changes in circulating immune evasive Omicron sub-variants. Despite these limitations, we studied a large population with nearly 8,000 breakthrough infection events during the Omicron era and were able to assess associations with key demographic and clinical characteristics driving SARS-CoV-2 acquisition using strong statistical methods.

## Conclusion

Our study sheds light on the complex interplay of factors influencing Omicron breakthrough infections in a large urban healthcare system in Chicago among a population of SARS-CoV-2 vaccinated and boosted individuals. We found a substantial one-year cumulative incidence of 5.1%, highlighting the ongoing challenges posed by SARS-CoV-2 despite vaccination efforts. These findings reflect real-world differences in immunity and exposure risk behaviors for populations vulnerable to Omicron variants of SARS-CoV-2 worldwide. By identifying key risk factors and disparities, our findings can inform targeted public health interventions to mitigate the impact of breakthrough infections in vaccinated populations. Public health messages should continue to emphasize the importance of considering both co-morbidities and age as critical factors in understanding and mitigating the risk of SARS-CoV-2 acquisition, ensuring that interventions are tailored to address the specific needs of vulnerable populations. As people make decisions about booster vaccinations, our findings provide more information for patients to consider personal risk in their decision making. Ongoing public health surveillance and research are crucial to understand the long-term effectiveness of vaccines against SARS-CoV-2. Future research should focus on understanding mechanisms of declining immunity, immune evasion by SARS-CoV-2 viruses, drivers of acquisition behavior, and optimizing protective vaccine.

## Supporting Information

**S1 Dataset. Vaccine Cohort Dataset.**

**S2 Supporting Information. Vaccine Cohort Dataset Codebook**

## Data Availability

All relevant data are within the manuscript and its Supporting Information files.

## References

1. Heath PT, Galiza EP, Baxter DN, Boffito M, Browne D, Burns F, et al. Safety and Efficacy of the NVX-CoV2373 COVID-19 Vaccine at Completion of the Placebo-Controlled Phase of a Randomized Controlled Trial. Clin Infect Dis. 2022. Epub 20221010. doi: 10.1093/cid/ciac803. PubMed PMID: 36210481.

2. Thomas SJ, Moreira ED, Jr., Kitchin N, Absalon J, Gurtman A, Lockhart S, et al. Safety and Efficacy of the BNT162b2 mRNA Covid-19 Vaccine through 6 Months. N Engl J Med. 2021;385(19):1761–73. Epub 20210915. doi: 10.1056/NEJMoa2110345. PubMed PMID: 34525277; PubMed Central PMCID: PMCPMC8461570.

3. Lewis NM, Self WH, Gaglani M, Ginde AA, Douin DJ, Keipp Talbot H, et al. Effectiveness of the Ad26.COV2.S (Johnson & Johnson) Coronavirus Disease 2019 (COVID-19) Vaccine for Preventing COVID-19 Hospitalizations and Progression to High Disease Severity in the United States. Clin Infect Dis. 2022;75(Supplement_2):S159–S66. doi: 10.1093/cid/ciac439. PubMed PMID: 35675695; PubMed Central PMCID: PMCPMC9214149.

4. Sahin U, Muik A, Derhovanessian E, Vogler I, Kranz LM, Vormehr M, et al. COVID-19 vaccine BNT162b1 elicits human antibody and TH1 T cell responses. Nature. 2020;586(7830):594-9. Epub 20200930. doi: 10.1038/s41586-020-2814-7. PubMed PMID: 32998157.

5. Haggenburg S, Lissenberg-Witte BI, van Binnendijk RS, den Hartog G, Bhoekhan MS, Haverkate NJE, et al. Quantitative analysis of mRNA-1273 COVID-19 vaccination response in immunocompromised adult hematology patients. Blood Adv. 2022;6(5):1537–46. doi: 10.1182/bloodadvances.2021006917. PubMed PMID: 35114690; PubMed Central PMCID: PMCPMC8816838.

6. Brosh-Nissimov T, Orenbuch-Harroch E, Chowers M, Elbaz M, Nesher L, Stein M, et al. BNT162b2 vaccine breakthrough: clinical characteristics of 152 fully vaccinated hospitalized COVID-19 patients in Israel. Clin Microbiol Infect. 2021;27(11):1652–7. Epub 20210707. doi: 10.1016/j.cmi.2021.06.036. PubMed PMID: 34245907; PubMed Central PMCID: PMCPMC8261136.

7. Islam JY, Vidot DC, Havanur A, Camacho-Rivera M. Preventive Behaviors and Mental Health-Related Symptoms Among Immunocompromised Adults During the COVID-19 Pandemic: An Analysis of the COVID Impact Survey. AIDS Res Hum Retroviruses. 2021;37(4):304–13. doi: 10.1089/AID.2020.0302. PubMed PMID: 33626959; PubMed Central PMCID: PMCPMC8035912.

8. Interim Clinical Considerations for Use of COVID-19 Vaccines in the United States: Center for Disease Control and Prevention 2023 [updated January, 2024]. Available from: https://www.cdc.gov/vaccines/covid-19/clinical-considerations/interim-considerations-us.html.

9. Starren JB, Winter AQ, Lloyd-Jones DM. Enabling a Learning Health System through a Unified Enterprise Data Warehouse: The Experience of the Northwestern University Clinical and Translational Sciences (NUCATS) Institute. Clin Transl Sci. 2015;8(4):269–71. Epub 20150601. doi: 10.1111/cts.12294. PubMed PMID: 26032246; PubMed Central PMCID: PMCPMC4553136.

10. Fox J. An R Companion to Applied Regression. Third ed. Thousand Oaks, CA: Sage Publications; 2019.

11. T T. A Package for Survival Analysis in R. R package version 3.5–5,2023.

12. Terry M. Therneau PMG. Modeling Survival Data : Extending the Cox Model. Springer, New York 2000.

13. Sjoberg DD FT. Competing Risks Estimation. R package version 0.2.0. 2022.

14. Stouten V, Hubin P, Haarhuis F, van Loenhout JAF, Billuart M, Brondeel R, et al. Incidence and Risk Factors of COVID-19 Vaccine Breakthrough Infections: A Prospective Cohort Study in Belgium. Viruses. 2022;14(4). Epub 20220413. doi: 10.3390/v14040802. PubMed PMID: 35458532; PubMed Central PMCID: PMCPMC9029338.

15. Sun J, Zheng Q, Madhira V, Olex AL, Anzalone AJ, Vinson A, et al. Association Between Immune Dysfunction and COVID-19 Breakthrough Infection After SARS-CoV-2 Vaccination in the US. JAMA Intern Med. 2022;182(2):153–62. doi: 10.1001/jamainternmed.2021.7024. PubMed PMID: 34962505; PubMed Central PMCID: PMCPMC8715386.

16. Gopinath S, Ishak A, Dhawan N, Poudel S, Shrestha PS, Singh P, et al. Characteristics of COVID-19 Breakthrough Infections among Vaccinated Individuals and Associated Risk Factors: A Systematic Review. Trop Med Infect Dis. 2022;7(5). Epub 20220522. doi: 10.3390/tropicalmed7050081. PubMed PMID: 35622708; PubMed Central PMCID: PMCPMC9144541.

17. COVID-19 Stats: COVID-19 Incidence,* by Urban-Rural Classification(dagger) - United States, January 22-October 31, 2020( section sign). MMWR Morb Mortal Wkly Rep. 2020;69(46):1753. Epub 20201120. doi: 10.15585/mmwr.mm6946a6. PubMed PMID: 33211682; PubMed Central PMCID: PMCPMC7676636.

18. Fazili A, Ain SN, Shah RJ, Raja FN, Farhat D, Nazir I. Incidence of breakthrough infections after COVID-19 vaccination among the COVID-19 vaccine recipients at a Tertiary Care Hospital in Srinagar. Indian J Public Health. 2023;67(2):305–8. doi: 10.4103/ijph.ijph_1403_22. PubMed PMID: 37459029.

19. Hacisuleyman E, Hale C, Saito Y, Blachere NE, Bergh M, Conlon EG, et al. Vaccine Breakthrough Infections with SARS-CoV-2 Variants. N Engl J Med. 2021;384(23):2212–8. Epub 20210421. doi: 10.1056/NEJMoa2105000. PubMed PMID: 33882219; PubMed Central PMCID: PMCPMC8117968.

20. Yazdanpanah M, Abadi B, Komendantova N, Zobeidi T, Sieber S. Some at Risk for COVID-19 Are Reluctant to Take Precautions, but Others Are Not: A Case From Rural in Southern Iran. Front Public Health. 2020;8:562300. Epub 20201116. doi: 10.3389/fpubh.2020.562300. PubMed PMID: 33304873; PubMed Central PMCID: PMCPMC7701237.

21. Kukreti S, Hsieh MT, Liu CH, Chen JS, Chen YJ, Hsieh MT, et al. Fear, Stress, Susceptibility, and Problematic Social Media Use Explain Motivation for COVID-19 Preventive Behaviors Among Patients With Stroke and Their Caregivers. Inquiry. 2024;61:469580231225030. doi: 10.1177/00469580231225030. PubMed PMID: 38314649; PubMed Central PMCID: PMCPMC10845975.

22. Mihelic A, Jelovcan L, Prislan K. Internal and external drivers for compliance with the COVID-19 preventive measures in Slovenia: The view from general deterrence and protection motivation. PLoS One. 2021;16(11):e0259675. Epub 20211115. doi: 10.1371/journal.pone.0259675. PubMed PMID: 34780530; PubMed Central PMCID: PMCPMC8592422.

23. Bashirian S, Jenabi E, Khazaei S, Barati M, Karimi-Shahanjarini A, Zareian S, et al. Factors associated with preventive behaviours of COVID-19 among hospital staff in Iran in 2020: an application of the Protection Motivation Theory. J Hosp Infect. 2020;105(3):430–3. Epub 20200428. doi: 10.1016/j.jhin.2020.04.035. PubMed PMID: 32360337; PubMed Central PMCID: PMCPMC7194681.

24. Smits PD, Gratzl S, Simonov M, Nachimuthu SK, Goodwin Cartwright BM, Wang MD, et al. Risk of COVID-19 breakthrough infection and hospitalization in individuals with comorbidities. Vaccine. 2023;41(15):2447–55. Epub 20230216. doi: 10.1016/j.vaccine.2023.02.038. PubMed PMID: 36803895; PubMed Central PMCID: PMCPMC9933320.

25. Walmsley S, Nabipoor M, Lovblom LE, Ravindran R, Colwill K, McGeer A, et al. Predictors of Breakthrough SARS-CoV-2 Infection after Vaccination. Vaccines (Basel). 2023;12(1). Epub 20231228. doi: 10.3390/vaccines12010036. PubMed PMID: 38250849; PubMed Central PMCID: PMCPMC10820583.

26. Willicombe M, Scanlon M, Loud F, Lightstone L. Should we be clinically assessing antibody responses to covid vaccines in immunocompromised people? BMJ. 2022;377:o966. Epub 20220412. doi: 10.1136/bmj.o966. PubMed PMID: 35414604.

27. Hansen CH, Michlmayr D, Gubbels SM, Molbak K, Ethelberg S. Assessment of protection against reinfection with SARS-CoV-2 among 4 million PCR-tested individuals in Denmark in 2020: a population-level observational study. Lancet. 2021;397(10280):1204–12. Epub 20210317. doi: 10.1016/S0140-6736(21)00575-4. PubMed PMID: 33743221; PubMed Central PMCID: PMCPMC7969130.

28. Liu C, Lee J, Ta C, Soroush A, Rogers JR, Kim JH, et al. Risk Factors Associated With SARS-CoV-2 Breakthrough Infections in Fully mRNA-Vaccinated Individuals: Retrospective Analysis. JMIR Public Health Surveill. 2022;8(5):e35311. Epub 20220524. doi: 10.2196/35311. PubMed PMID: 35486806; PubMed Central PMCID: PMCPMC9132195.

29. White EM, Yang X, Blackman C, Feifer RA, Gravenstein S, Mor V. Incident SARS-CoV-2 Infection among mRNA-Vaccinated and Unvaccinated Nursing Home Residents. N Engl J Med. 2021;385(5):474–6. Epub 20210519. doi: 10.1056/NEJMc2104849. PubMed PMID: 34010526; PubMed Central PMCID: PMCPMC8174028.

30. O’Rourke K. Age and smoking predict antibody titers after the BNT162b2 COVID-19 vaccine. Cancer. 2022;128(3):431. doi: 10.1002/cncr.34082. PubMed PMID: 35050525; PubMed Central PMCID: PMCPMC9015587.

31. Bartleson JM, Radenkovic D, Covarrubias AJ, Furman D, Winer DA, Verdin E. SARS-CoV-2, COVID-19 and the Ageing Immune System. Nat Aging. 2021;1(9):769–82. Epub 20210914. doi: 10.1038/s43587-021-00114-7. PubMed PMID: 34746804; PubMed Central PMCID: PMCPMC8570568.

32. Blomberg BB, Frasca D. Quantity, not quality, of antibody response decreased in the elderly. J Clin Invest. 2011;121(8):2981–3. Epub 20110725. doi: 10.1172/JCI58406. PubMed PMID: 21785210; PubMed Central PMCID: PMCPMC3148749.

33. CDC. nterim Clinical Considerations for Use of COVID-19 Vaccines in the United States Centers for Disease and Control Prevention2024. Available from: https://www.cdc.gov/vaccines/covid-19/clinical-considerations/interim-considerations-us.html#:~:text=Summary%20of%20recent%20changes%20(last,Novavax%2C%20Pfizer%2DBioNTech).C.

34. Dash NR, Barqawi HJ, Obaideen AA, Al Chame HQ, Samara KA, Qadri R, et al. COVID-19 Breakthrough Infection Among Vaccinated Population in the United Arab Emirates. J Epidemiol Glob Health. 2023;13(1):67–90. Epub 20230216. doi: 10.1007/s44197-023-00090-8. PubMed PMID: 36795274; PubMed Central PMCID: PMCPMC9933808.

35. Wilson RF, Sharma AJ, Schluechtermann S, Currie DW, Mangan J, Kaplan B, et al. Factors Influencing Risk for COVID-19 Exposure Among Young Adults Aged 18-23 Years - Winnebago County, Wisconsin, March-July 2020. MMWR Morb Mortal Wkly Rep. 2020;69(41):1497–502. Epub 20201016. doi: 10.15585/mmwr.mm6941e2. PubMed PMID: 33056953; PubMed Central PMCID: PMCPMC7561092.

36. Rolison JJ, Hanoch Y, Wood S, Liu PJ. Risk-taking differences across the adult life span: a question of age and domain. J Gerontol B Psychol Sci Soc Sci. 2014;69(6):870–80. Epub 20131022. doi: 10.1093/geronb/gbt081. PubMed PMID: 24149517.

37. Malhotra S, Mani K, Lodha R, Bakhshi S, Mathur VP, Gupta P, et al. SARS-CoV-2 Reinfection Rate and Estimated Effectiveness of the Inactivated Whole Virion Vaccine BBV152 Against Reinfection Among Health Care Workers in New Delhi, India. JAMA Netw Open. 2022;5(1):e2142210. Epub 20220104. doi: 10.1001/jamanetworkopen.2021.42210. PubMed PMID: 34994793; PubMed Central PMCID: PMCPMC8742193.

38. de Gier B, de Oliveira Bressane Lima P, van Gaalen RD, de Boer PT, Alblas J, Ruijten M, et al. Occupation- and age-associated risk of SARS-CoV-2 test positivity, the Netherlands, June to October 2020. Euro Surveill. 2020;25(50). doi: 10.2807/1560-7917.ES.2020.25.50.2001884. PubMed PMID: 33334396; PubMed Central PMCID: PMCPMC7812419.

39. Rumain B, Schneiderman M, Geliebter A. Prevalence of COVID-19 in adolescents and youth compared with older adults in states experiencing surges. PLoS One. 2021;16(3):e0242587. Epub 20210310. doi: 10.1371/journal.pone.0242587. PubMed PMID: 33690600; PubMed Central PMCID: PMCPMC7946189.

40. Schneiderman M, Rumain B, Kaganovskiy L, Geliebter A. Incidence and Relative Risk of COVID-19 in Adolescents and Youth Compared With Older Adults in 19 US States, Fall 2020. JAMA Netw Open. 2022;5(7):e2222126. Epub 20220701. doi: 10.1001/jamanetworkopen.2022.22126. PubMed PMID: 35838670; PubMed Central PMCID: PMCPMC9287748.

41. McKeigue PM, McAllister DA, Hutchinson SJ, Robertson C, Stockton D, Colhoun HM. Vaccine efficacy against severe COVID-19 in relation to delta variant (B.1.617.2) and time since second dose in patients in Scotland (REACT-SCOT): a case-control study. Lancet Respir Med. 2022;10(6):566–72. Epub 20220225. doi: 10.1016/S2213-2600(22)00045-5. PubMed PMID: 35227416; PubMed Central PMCID: PMCPMC8880999.

42. Lim SY, Kim JY, Jung J, Yun SC, Kim SH. Waning of humoral immunity depending on the types of COVID-19 vaccine. Infect Dis (Lond). 2023;55(3):216–20. Epub 20230110. doi: 10.1080/23744235.2023.2165707. PubMed PMID: 36625442.

43. Andrews N, Stowe J, Kirsebom F, Toffa S, Rickeard T, Gallagher E, et al. Covid-19 Vaccine Effectiveness against the Omicron (B.1.1.529) Variant. N Engl J Med. 2022;386(16):1532–46. Epub 20220302. doi: 10.1056/NEJMoa2119451. PubMed PMID: 35249272; PubMed Central PMCID: PMCPMC8908811.

44. Tan WC, Tan JYJ, Lim JSJ, Tan RYC, Lee A, Leong FL, et al. COVID-19 Severity and Waning Immunity After up to 4 mRNA Vaccine Doses in 73 608 Patients With Cancer and 621 475 Matched Controls in Singapore: A Nationwide Cohort Study. JAMA Oncol. 2023;9(9):1221–9. doi: 10.1001/jamaoncol.2023.2271. PubMed PMID: 37440245; PubMed Central PMCID: PMCPMC10346511.

45. Intawong K, Chariyalertsak S, Chalom K, Wonghirundecha T, Kowatcharakul W, Thongprachum A, et al. Waning vaccine response to severe COVID-19 outcomes during omicron predominance in Thailand. PLoS One. 2023;18(5):e0284130. Epub 20230511. doi: 10.1371/journal.pone.0284130. PubMed PMID: 37167215; PubMed Central PMCID: PMCPMC10174527.

46. Feng A, Obolski U, Stone L, He D. Modelling COVID-19 vaccine breakthrough infections in highly vaccinated Israel-The effects of waning immunity and third vaccination dose. PLOS Glob Public Health. 2022;2(11):e0001211. Epub 20221109. doi: 10.1371/journal.pgph.0001211. PubMed PMID: 36962648; PubMed Central PMCID: PMCPMC10021336.

47. Khanijahani A, Iezadi S, Gholipour K, Azami-Aghdash S, Naghibi D. A systematic review of racial/ethnic and socioeconomic disparities in COVID-19. Int J Equity Health. 2021;20(1):248. Epub 20211124. doi: 10.1186/s12939-021-01582-4. PubMed PMID: 34819081; PubMed Central PMCID: PMCPMC8611382.

48. Spruce L. Back to Basics: Social Determinants of Health. AORN J. 2019;110(1):60–9. doi: 10.1002/aorn.12722. PubMed PMID: 31246307.

49. Abrams EM, Szefler SJ. COVID-19 and the impact of social determinants of health. Lancet Respir Med. 2020;8(7):659–61. Epub 20200518. doi: 10.1016/S2213-2600(20)30234-4. PubMed PMID: 32437646; PubMed Central PMCID: PMCPMC7234789.

50. Sun J, Patel RC, Zheng Q, Madhira V, Olex AL, Islam JY, et al. COVID-19 Disease Severity among People with HIV Infection or Solid Organ Transplant in the United States: A Nationally-representative, Multicenter, Observational Cohort Study. medRxiv. 2021. Epub 20210728. doi: 10.1101/2021.07.26.21261028. PubMed PMID: 34341798; PubMed Central PMCID: PMCPMC8328066.

51. Agyemang C, Richters A, Jolani S, Hendriks S, Zalpuri S, Yu E, et al. Ethnic minority status as social determinant for COVID-19 infection, hospitalisation, severity, ICU admission and deaths in the early phase of the pandemic: a meta-analysis. BMJ Glob Health. 2021;6(11). doi: 10.1136/bmjgh-2021-007433. PubMed PMID: 34740916; PubMed Central PMCID: PMCPMC8573300.

52. Lopez L, 3rd, Hart LH, 3rd, Katz MH. Racial and Ethnic Health Disparities Related to COVID-19. JAMA. 2021;325(8):719–20. doi: 10.1001/jama.2020.26443. PubMed PMID: 33480972.

